# A Precision Medicine Framework for Personalized Simulation of Hemodynamics in Cerebrovascular Disease

**DOI:** 10.1101/2020.01.28.20019190

**Authors:** Dietmar Frey, Michelle Livne, Heiko Leppin, Ela M Akay, Orhun U Aydin, Jonas Behland, Jan Sobesky, Peter Vajkoczy, Vince I Madai

**Affiliations:** Charité Lab for Artificial Intelligence in Medicine, Department of Neurosurgery, Charité University Medicine Berlin, Berlin, Germany; Johanna Etienne Hospital Neuss, Germany; Centre for Stroke Research Berlin, Charité University Medicine Berlin, Berlin, Germany; Department of Neurosurgery, Charité University Medicine Berlin, Berlin, Germany

## Abstract

**Introduction:** Cerebrovascular disease is a major public health challenge. An important biomarker is cerebral hemodynamics. To measure cerebral hemodynamics, however, only invasive, potentially harmful or time-to-treatment prolonging methods are available. We present a simulation-based alternative which allows calculation of cerebral hemodynamics based on the individual vessel configuration of a patient derived from structural vessel imaging.

**Methods:** We implemented a framework allowing annotation of extracted brain vessels from structural imaging followed by 0-dimensional lumped modelling of cerebral hemodynamics. For annotation, a 3D-graphical user interface (GUI) was implemented. For 0D-simulation, we used a modified nodal analysis (MNA), which was adapted for easy implementation by code. The code was written in-house in java. The simulation GUI allows inspection of simulation results, identification of vulnerable areas, simulation of changes due to different systemic blood pressures. Moreover, sensitivity analysis was implemented allowing the live simulation of changes of variables such as vessel lumen to simulate procedures and disease courses. In two exemplary patients, simulation results were compared to dynamic-susceptibility-weighted-contrast-enhanced magnetic-resonance(DSC-MR) perfusion imaging.

**Results:** The successful implementation of the framework allowing individualized annotation and simulation of patients is presented. In two exemplary patients, both the simulation as well as DSC-MRI showed the same results pertaining to the identification of areas vulnerable to ischemia. Sensitivity analysis allows the individualized simulation of changes in vessel lumen and the effect on hemodynamics.

**Discussion:** We present the first precision medicine pipeline for cerebrovascular disease which allows annotation of the arterial vasculature derived from structural vessel imaging followed by personalized simulation of brain hemodynamics. This paves the way for further development of precision medicine in stroke using novel biomarkers and might make the application of harmful and complex perfusion methods obsolete for certain use cases in the future.

## Introduction

Cerebrovascular disease, and in particular stroke, is a major public health challenge. It is a leading cause of death and disability worldwide(“WHO EMRO | Stroke, Cerebrovascular Accident | Health Topics” 2019). While there have been advances in prevention and treatment in the past – e.g. mechanical thrombectomy for acute stroke treatment - the overall prevention and treatment results still remain poor(Benjamin et al. 2019). A potential game-changer of stroke treatment success is precision medicine(Hinman et al. 2017; Rostanski and Marshall 2016). It aims to provide personalized therapy recommendations based on the individual features of the patient. It utilizes today’s plethora of available patient data as well as mathematical modeling to offer individualized predictions for patients(Hinman et al. 2017; Rostanski and Marshall 2016). While highly promising, precision medicine relies on the presence of informative data allowing the differentiation of pathology patterns(Hinman et al. 2017; Livne 2019). In cerebrovascular disease, important information about the severity of stroke risk and potential response to treatment is encoded in individual pathophysiological parameters - in biomarkers - which can be recorded to aid decision making. Here, one of the most important is the hemodynamic status(Sobesky 2012; Campbell et al. 2019). This biomarker is already used in a precision medicine approach to identify individual patients benefiting from thrombolysis beyond the currently established treatment time windows which is crucial since often treatment is denied due time constraints (Campbell et al. 2019). In chronic cerebrovascular disease, it might aid by identifying areas which are highly vulnerable to stroke(Mutke et al. 2014; Martin et al. 2015). In the clinical setting, however, this data is only available using specialized methodologies, i.e. Dynamic Susceptibility-weighted Contrast-enhanced Magnetic Resonance Imaging (DSC-MRI) perfusion, Computed-Tomography (CT)-perfusion, Arterial Spin Labeling (ASL) perfusion or functional MRI(Sobesky 2012; Max Wintermark et al. 2013; M Wintermark et al. 2005; Kamalian and Lev 2019; Okell et al. 2019; Khalil et al. 2018). These techniques may harm patients through contrast agents, significantly prolong the time to treatment and lead to increased costs. Also, standardization of these complex methods is highly challenging(Sobesky 2012; Calamante 2013; Zaharchuk 2011).

An alternative approach to derive biomarkers for precision medicine is the transformation of routinely acquired data by mechanistic simulations(Fröhlich et al. 2018). These simulations integrate domain knowledge by mathematically describing known disease-driving core processes(Fröhlich et al. 2018). Interestingly for cerebrovascular disease, several works in the past have developed general mechanistic simulations of the blood flow in the brain, e.g. Alastruey et al. and Grinberg et al.(Alastruey et al. 2007; Grinberg et al. 2009). These simulations have the potential to become a contrast-agent free biomarker of hemodynamics for the diagnosis and treatment of cerebrovascular diseases. However, for these simulations personalization on an individual patient level is still pending making it not applicable in a clinical setting.

Thus, the novel idea presented in this work is a software framework to transform routine structural vessel imaging data as an input to a mechanistic simulation of individual hemodynamics for a given patient. The unique vessel configuration of each patient can be used to simulate hemodynamics to potentially identify areas that are vulnerable in case of stenosis and occlusion. Several use cases can be envisioned for such a framework. It could allow assessment of stroke risk, pre-operative simulation of interventional success like thrombectomy in acute stroke, preventive or therapeutic endarterectomy and stenting of brain-supplying vessels, respectively. Another highly interesting, if rather rare case is the simulation of the outcome of extracranial-intracranial (EC-IC) bypass surgery, e.g. in Moya-Moya disease. Here, there is a special need to predict the success of the surgery(Wessels, Hecht, and Vajkoczy 2019). Lastly, the simulation information could be used for the prediction of stroke outcome in conjunction with other clinical and imaging parameters enabling clinicians with an objective criterion for decision support in the acute setting.

Thus, the objective of the presented work was to provide a framework allowing the incorporation of individual structural vessel data to simulate areas of higher hemodynamic vulnerability as a disease biomarker. For this purpose, we developed a pipeline consisting of the following sequential steps: 1) Segmentation of vessel information from structural data, in our case from time-of-flight (TOF) magnetic resonance imaging (MRI). 2) Annotation of the vessel tree with an easy-to-use graphical user interface (GUI). And 3) Simulation where results can be inspected and different blood pressure scenarios can be simulated by the user.

The simulation was implemented as a steady-state zero-dimensional lumped model of the Circle-of-Willis (CoW) and major brain artery circulation. We included individual vessel resistances by 1-dimensional calculation using the individual length and the width of the arteries from patient structural vessel imaging. Hemodynamic measures were calculated using an adapted version of the modified nodal analysis (MNA)((Chung-Wen Ho, Ruehli, and Brennan 1975) coined AMNA, where we simplify the solution of the matrix equations facilitating easier implementation and faster runtime. We show the implemented framework in detail - including a video of the annotation process - as well as two exemplary simulation results of patients with cerebrovascular disease in comparison with perfusion imaging.

## Methods

### Patients

Two patients with steno-occlusive disease from an imaging study of cerebral perfusion in stroke patients (PEGASUS study(Mutke et al. 2014; Martin et al. 2015)) were evaluated as examples for the presented framework. This study was approved by the institutional ethics committee of Charité Universitätsmedizin Berlin and the patients gave written informed consent. One patient with MCA stenosis and one patient with carotid artery occlusion were chosen. The patients were randomly chosen from all patients with the aforementioned vessel conditions.

### The framework pipeline

The framework pipeline consists of these chronological steps: a) Segmentation, b) Skeletonization, c) Annotation, and d) Simulation. In the following, each of the steps is described in detail.

### Segmentation

Structural MRI imaging consisted of Time-of-Flight(TOF)-MR images. The images were first segmented using a neural net segmentation model developed in our group(Livne et al. 2019). The segmentations were then manually corrected to derive ground-truths standards. The segmentations are voxel-based binary representations of the vessel tree for each individual patient. The imaging parameters for TOF-MR-images in the PEGASUS study were: voxel size = (0.5 × 0.5 × 0.7) mm3; matrix size: 312 × 384 × 127; TR/TE = 22 ms/3.86ms; time of acquisition: 3:50min, flip angle = 18 degrees.

### Skeletonization

The segmentations were skeletonized using the DtfSkeletonization module(“DtfSkeletonization — MeVisLab Documentation” n.d.) of MeVisLab(“Website: MeVisLab” n.d.). Here, a one voxel skeleton of the vessel midpoints is created with the radius encoded in the voxel value. This skeleton volume is then transferred to the manual annotation module.

### Annotation

The annotation module was developed and coded from scratch by the in-house development team.

Within the annotation framework, the transferred skeleton volume is transformed into a Java-based tree structure - the so-called skeleton graph - representing the skeleton as a set of edges containing all necessary geometric information for 3D rendering, and a set of junction vertices. This tree structure is loaded via a RESTful service interface in JSON format into a JavaScript-based web frontend where it is rendered by using the Three.js library as a rotatable and zoomable 3D view (see figure 8).

Within the 3D view, it is possible to select edges and tag them with an item from a list of vessel (artery) descriptors and their anatomic location (visualized by color) interactively. The triple consisting of an edge, a tagged vessel item, and an anatomic location is defined as an annotation. A set of made annotations can be saved in the backend via a RESTful service interface within an appropriate Java presentation. In the final step, based on a skeleton graph and a belonging annotation set, a simulation model serving as the input for our simulation component can be created once the annotation is finished.

Within our framework it is possible to annotate also 3rd order branches (A3, M3, P3). For exemplary patients in our study, annotations were performed until 2nd order branches (A2, M2 and P2), since higher order vessels are unlikely to play a crucial role in steno-occlusive disease.

### Simulation

The simulation was developed and coded from scratch in-house in collaboration with the Zuse Institute Berlin of the Free University Berlin(“ZIB | Zuse Institute Berlin (ZIB)” n.d.). Our model describes the cerebral vascular tree by a planar graph, which is given in figure 1. The blood flow through the vessel tree is modeled in analogy to electric circuits in a modified nodal analysis.

**Figure 1:**
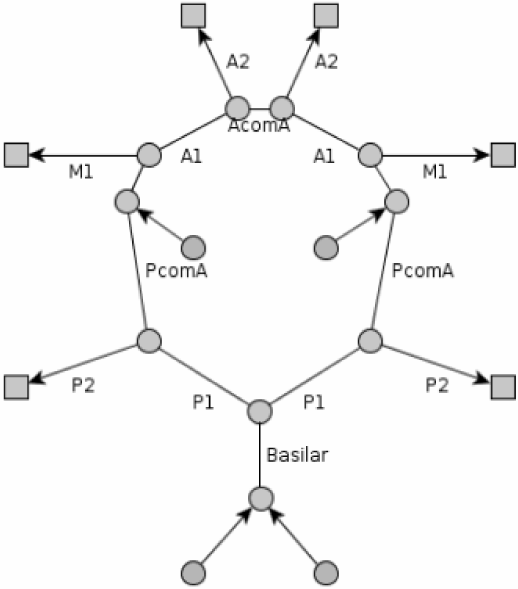
The circle of Willis represented by edges, nodes and supply areas‥ The alphanumeric abbreviations and AcomA and PcomA stand for certain brain vessels (see legend) represented by edges. The round shapes represent nodes, whereas the quadratic shapes represent supply areas. *M1: A1: anterior cerbral artery (ACA) segment 1, A2: ACA segment 2, M1: middle cerbral artery (MCA) segment 1, P1: posterior cerbral artery(PCA) segment 1, P2: PCA segment 2, PcomA and AcomA: posterior and anterior communicating arteries*.

**Figure 2:**
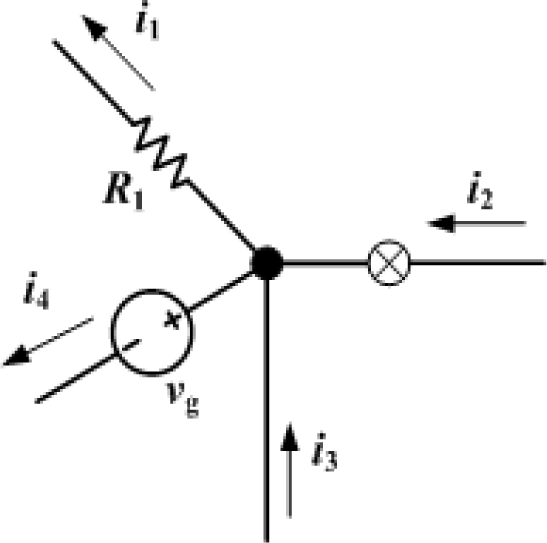
Graphical illustration Kirchhoff’s first law. The current entering a node = *i*_2_ + *i*_3_ must equal the current leaving that node =*i*_1_ + *i*_4_. See blood flow analogy equation 5 for the Mass flow law. In the figure i correspond to current, R represents resistance and v stands for the voltage.

Edges in the graph represent blood vessels, while nodes represent either supply areas, blood sources or junctions between nodes. Supply areas are marked with square-shaped nodes, initial (round) nodes are blood sources and the rest of the (round) nodes are junctions. The arrows of the edges indicate the flow directions to the supply areas or away from source nodes. The blood flow into the supply areas is provided by the outgoing segments (A2, P2 and M1 or M2) of the circle of Willis.

#### Modeling the flow and vessel network

The cerebral vessel tree retains an overall Reynolds number allowing to describe the cerebral blood flow in terms of a Newtonian fluid. The arteries are modeled as perfect cylinders and the drop in blood pressure *ΔP* along a cerebral artery of length *L*, with radius *r*, blood dynamic viscosity μ for a volumetric flow rate *Q* is determined according to Hagen-Poiseuille equation:

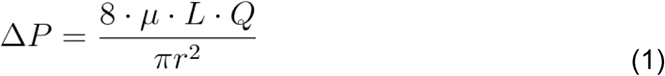

Hagen-Poiseuille equation is equivalent to Ohm’s law. Therefore, the resistance of an arterial vessel can be defined as:

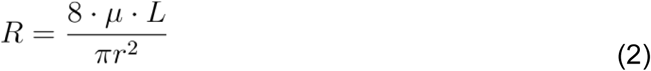

In the presented use case the vessel diameter is not homogeneous across the whole vessel segment and consequently equation 2 is invalid. To account for variable diameters over a segment, the fluid dynamics is applied on infinitely small segments with a constant radius to yield the equation for non-constant radii. The resistance can be therefore derived using the following integral equation:

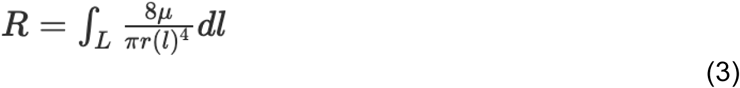

In practice, discrete application is used, as a segment is defined by a diameters-vector of length n-1 = the number of voxels in the segment. To account for the fact that an antiderivative can only be determined for segments that can be constantly differentiated, we need to approximate the condition of continuity. For this purpose, the radius *r*(*l*) is described by a continuous linear extrapolation function, which connects the radius of a given voxel with the next voxel in a linear fashion to allow the calculation of (3):

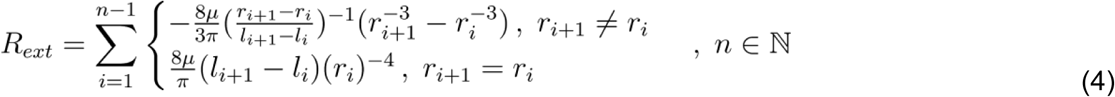

Where *R*_*ext*_ stands for the extrapolated resistance. For the full derivation of *R*_*ext*_ see appendix 1. According to the mass flow law, the amount of blood entering a node must equal the amount of blood that leaves a node. Mathematically this is described as follows:

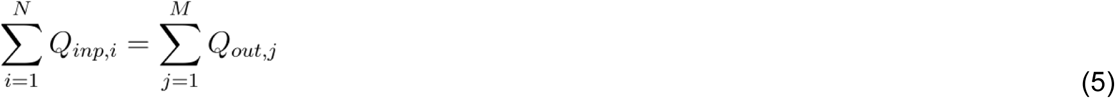

Where *Q*_*inp,i*_ is the input blood flow from source i to node n, for N input sources, and *Q*_*inp,i*_ is the output from node n through vessel j, for M outputs.

The mass flow conservation can be then applied using this equation for each node and the resulting set of equations can be solved to yield the pressures. In the presented application, it is applied via a Modified Nodal Analysis (MNA) (Chung-Wen Ho, Ruehli, and Brennan 1975) that incorporates constraints on the system to be driven by a system pressure and ensures constant blood supply to specific regions.

Our network consists of three types of nodes and two types of edges. The primary type of node is one that connects different vessels with each other (i.e. junctions between vessels). The second type of node is a source that provides the system with blood. The third type of node is a supply area, whose resistance depends on the incident pressure and the auto-regulation process described in the next section (see figure 1). The types of edges in our network represent normal arterial blood vessels and vessels that connect the supply areas to the sink node. The resistance of the latter is determined via the auto-regulation function given in the next section.

#### Modelling auto-regulation

The simulated vessel network encompasses the circle of Willis and the larger arterial segments of 1st and 2nd order of the three major brain arteries: anterior-, medial- and posterior cerebral artery (ACA, MCA and PCA). Those are the vasculature segments that are a) accessible for interventions such as surgery or thrombectomy and b) their anatomical architecture can be derived from medical imaging. The vascular downstream regions after the above segments including the 3rd order segments represent a network of small arteries, finer small arterioles and the capillary bed that can change their radii in order to decrease or increase the blood supply. This process is called auto-regulation and ensures that the blood supply to the brain remains largely constant within certain limits. Most of the vascular network’s resistance originates from these supply areas. Autoregulation was implemented into the simulation framework according to the following equation based on literature values(Lang et al. 2005):

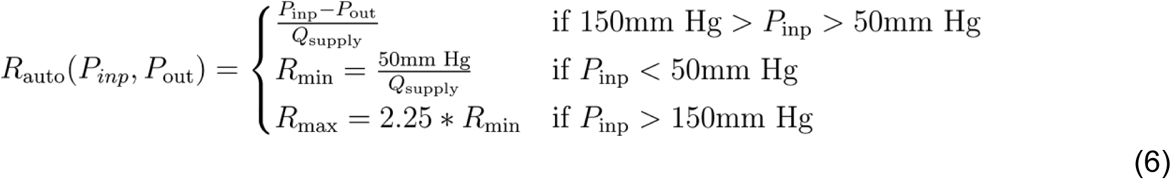

Where *R*_*auto*_is the autoregulated resistance of the vessel, *P*_*inp*_ and *P*_*out*_are the input and output blood pressures of the vessel and *Q*_*supply*_ is the blood flow supply to the vessel.

In more detail, the blood flow into the supply areas is provided by the outgoing segments (A2, P2 and M1 or M2) of the circle of Willis, see figure 1. The behavior of the supply areas is modeled according to the autoregulation as described in equation 6. This means that the peripheral resistance of the supply area adjusts itself such that the blood flow is kept constant for a given pressure gradient *ΔP*.

#### Boundary Conditions

The model requires a systemic mean arterial pressure (MAP) that drives the flow through the network such that the blood supply to the various supply areas of the brain is constant, while the blood flows and pressures are adjusted accordingly. However, the auto-regulation model is limited to the region between 80mmHg and 180mmHg. Blood pressure below or above these boundaries indicates a pathological state in which the body cannot maintain the necessary blood pressure to ensure constant blood supply and results in hypoperfusion of the brain tissue. Under these conditions, the simulation indicates that the respective supply areas are not sufficiently supplied with blood. Clinically speaking, these areas are vulnerable to ischemia. In addition to the system pressure a blood flow rate per supply area was provided (see table 1).

**Table 1:** Boundary Conditions.

| Symbol | Physical Quantity |
| --- | --- |
| $P_{in}$ | System pressure |
| $Q_{s,i}$ for $i=\{1,...,N\}$ | Blood supply demand of supply area i |

#### Integration of Physiological Data

The physiological properties of the vessels length and radius that were used to generate a default normal vessel tree in the absence of individual patient data can be found in appendix 3. When patient data is imported the default values are replaced by the individual patient data.

#### Algorithmic derivation of the blood supply

The blood supply to the specified brain areas is derived using an adjusted Modified Nodal Analysis (AMNA). The construction of the matrix equations per type of node and edge is detailed in the following. The described system is overdetermined by N equations, where N stands for the number of nodes. The last node is taken as the sink node, with a pressure value of 0. Therefore, the system is described by N-1 mass flow equations. These mass flow equations can be written in terms of a matrix:

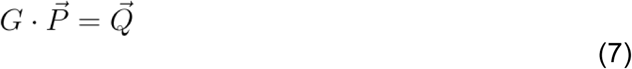

Where the matrix *G* represents the network structure and consists of the conductivity (e.g. inverse resistances) of the vessels. 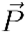 is the unknown blood pressure vector and 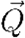 is the blood flow vector. The AMNA algorithm yields a reduced size of the solution system by removal of known values from the solution vector 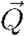. The following section details the determination of the conductivity values and blood flow to construct the matrix equations from a vessel graph as depicted in figure 1 and followed by the AMNA implementation.

### Junction nodes

The matrix row vector for a junction node is defined as:

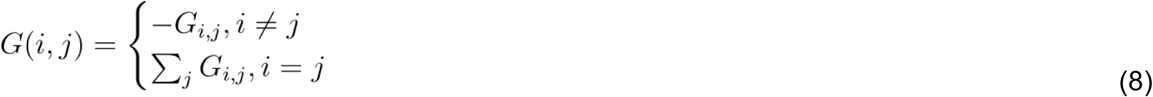

where the diagonal element is the sum of all *G* values for all connected edges and the non-diagonal elements are set to be the negative conductivity value of the corresponding edge - or otherwise 0. An exemplary derivation of a junction node vector is described under figure 4. The corresponding element of the 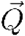 is 0. This represents a flow equation as depicted by equation 5.

**Figure 4:**
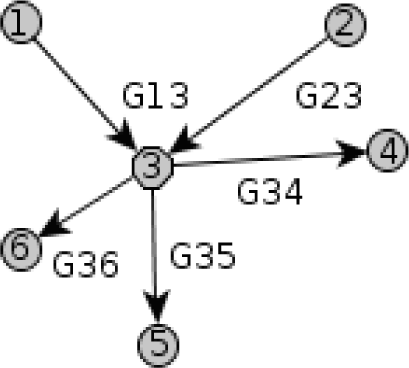
Illustration of a normal node representing a vessel junction. Each numbered circle in the figure represents a node and each arrow represents a directed edge. The figure illustrates a normal node - numbered as 3 - with two incident edges (G13 and G23) and three outgoing edges (G34, G35 and G36). The resulted G matrix row vector for node 3 is: *G*(3,3) = *G*_3,1_ + *G*_3,2_ + *G*_3,4_ + *G*_3,5_ + *G*_3,6_, 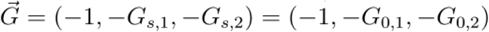

### Source nodes

The matrix row vector for a source node is defined as:

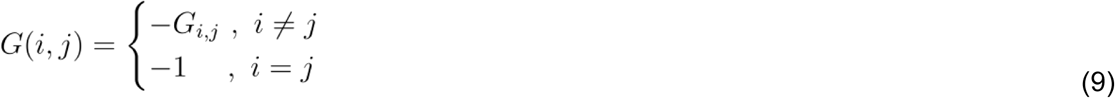

The diagonal element is *G*(*i,i*) = −1 and all other matrix elements are the negative conductivity values of the (connected) corresponding edges and otherwise 0, similarly to junction nodes. An exemplary derivation of the vector is described in figure 5. The flow into the source node is determined via the supply areas and according to the mass flow can be written as:

**Figure 5:**
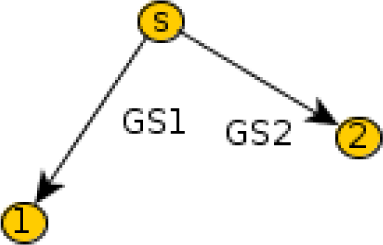
Illustration of a source node. Each numbered circle in the figure represents a node and each arrow represents a directed edge. The figure illustrates a graph segment with a source node (marked as s) and two vessels (GS1 and GS2). The resulted G matrix row vector for node s = 0 is: 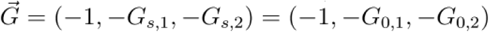, which results in the overall Nodal equation:*qs =* −(*G*_s,1_ + *G*_s,2_)*P*_0_

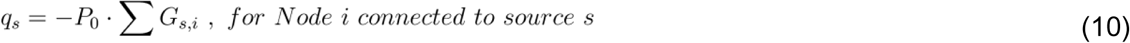

For the nodes that are directly connected with the source node, the q vector element is then:

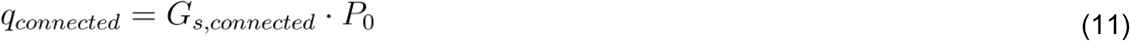

Figure 5 depicts a graph segment with a source node and two connected vessels.

### Supply Nodes

To model supply areas, the auto-regulation equation is applied (equation 6). The matrix row vector for a supply node is defined as:

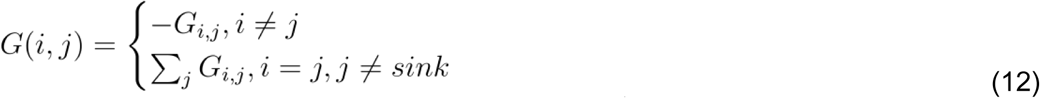

Similarly to junction nodes, the diagonal element *G*(*i,i*) is the conductivity sum of incident edges, however in case the edge is connected to a sink it does not contribute to the sum. An exemplary derivation of the vector is described in figure 6. The q-vector values are:

*q*_*i*_ = − *Q*_*s,i*_ for the supply node i connected to source s.

And *q*_*sink*_ = *Q*_*s,i*_ for the sink connected to supply node i.

Figure 6 depicts a case with two supply areas, denoted by the thick dashed lines that are connected to the same sink.

**Figure 6:**
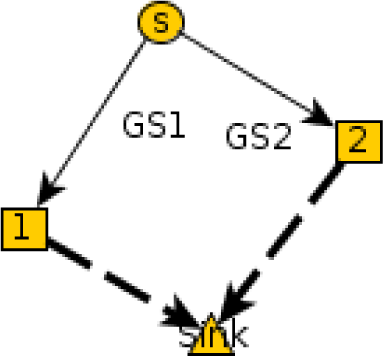
Illustration of a supply node. Each numbered circle or square in the figure represents a node and each arrow represents a directed edge. Dashed edges correspond to supply areas that are governed by auto-regulation. The figure illustrates two supply node - numbered 1 and 2. Sink represents the analogy to current sink.The resulted G matrix row vector for the supply node 1 is:

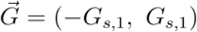

Which results in the overall Nodal equation:

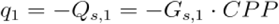

Where CPP stands for cerebral perfusion pressure

### Summary of nodal analysis construction

To summarize, the q-vector has a contribution for a supply area or a source node. If an edge connects a source with a supply area, the corresponding q-vectors will have contributions from both the source and the supply area. Table 3 summarizes the terms for the q-vector elements in dependence of the node type.

**Table 3:** Summary of q vector element derivation in the modified nodal analysis (MNA).

| Type of Node | Q-vector Node | Q-vector Incident Node |
| --- | --- | --- |
| Source | $q_s = -P_0 \cdot \sum G_{s,i}$ | $G_{s,connected} \cdot P_0$ |
| Supply Area | $-Q_{s,is}$ | $Q_{s,is}$ |
| Junction | 0 | 0 |

Table 4 summarizes the algorithm for the creation and determination of the G-matrix.

**Table 4:** Summary of G-matrix i-th row elements determination in the modified nodal analysis (MNA).

| Element | Value |
| --- | --- |
| $G(i, i)$ | $\sum_{i \neq j}^N G_{i,j}$ |
| $G(i, j)$ | $-G_{i,j}$ |

The edge conductivity is described by equation (13). Here, the edge conductivity between a supply node and the sink is determined by the auto-regulation function.

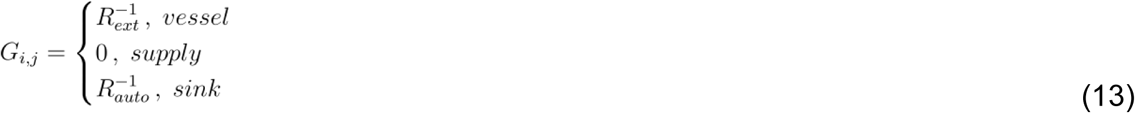

Here vessel refers to a connection of junction-node to junction-node, supply refers to a connection of junction node to a supply node and sink refers to a connection of a supply node to sink. *R*_*ext*_ and *R*_*auto*_ are described in formulas 4 and 6 respectively.

#### Adjusted Modified Nodal Analysis (AMNA)

The AMNA allows to reduce the size of the solution system by removing known values from the solution vector 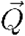 in consecutive 3 steps. Once the MNA matrix equations are determined, all the values of the system matrix that are known given system pressures are drawn to the right side solution vector 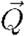. This pertains the values associated with the input pressures of the source nodes, i.e. MAP. As a result of this transition the equations associated with the source nodes become zero. In a second step, these redundant rows and columns are then deleted. Finally the G matrix columns are swapped to yield a diagonal matrix, with the solution vector adjusted accordingly. This process simplifies the solution derivation and therefore accelerates its application.

#### Sensitivity Analysis

In order to predict the effects of changes of a certain variable on the system, in this case the radius of a blood vessel, we performed a sensitivity analysis. Sensitivity analysis quantifies this effect by estimating the partial derivative of a system variable such as the blood pressure in this case, with respect to the radius of a given vessel. This is achieved by application of Newton-Raphson method using Taylor series expansion as follows(Gopalakrishna and Greimann 1988):

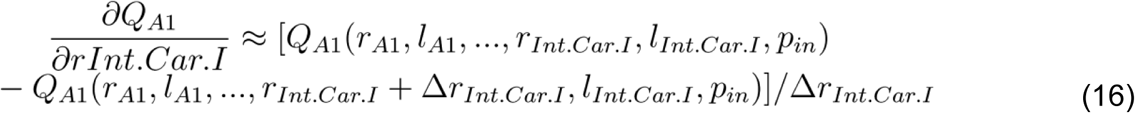

Similar Newton steps can be formulated for all relevant system variables and allow to estimate how the system reacts to changes of certain system variables.

### Simulation interface

The simulation interface was implemented as a java application with an integrated graphical user interface (GUI) under the loose-coupling paradigm to ensure that components can be exchanged easily. The key element of the simulation is a 2D projection as a representation of the simulated vessel tree. The interface’s main components are the simulation view including areas at risk and pressure view, and the possibility to change blood pressure as a boundary condition.

### Perfusion Imaging processing

DSC imaging was processed using the pgui software (Version 1.0, Center for functional neuroimaging, Aarhus University). Four arterial input functions were placed contralateral to the stenosis/occlusion in the M2 vessel area and visually assessed for optimal shape(Calamante 2013; 2013). Deconvolution was performed according to the parametric method introduced by Mouridsen et al(Mouridsen et al. 2014). Non-deconvolved time-to-peak, and deconvolved cerebral blood flow(CBF), time-to-maximum (Tmax) and mean-transit-time(MTT) maps were created and assessed in this study.

### Comparison of simulation and perfusion imaging results

In both exemplary patients simulation results were obtained for normal blood pressure (120/80) as well as for low blood pressure (90/60). The simulation results were inspected for areas at risk denoted by red areas equaling critically low perfusion pressure. At the same time DSC maps were inspected for signs of hemodynamic changes indicating tissues at risk, such as increased TTP, low CBF, increased Tmax and increased MTT.

## Results

### The framework

Figure 7 gives an overview of the steps of the framework. The input is a binary vessel (Figure 7B) mask segmented from a structural TOF-image(Figure 7A). This image is then skeletonized (Figure 7C). This skeleton can be imported in our annotation tool, where the vessel tree is displayed in a 3D fashion and annotation is performed (Figure 7D and 7E). The annotated vasculature can then be transferred to the simulation tool (Figure 7F).

**Figure 7:**
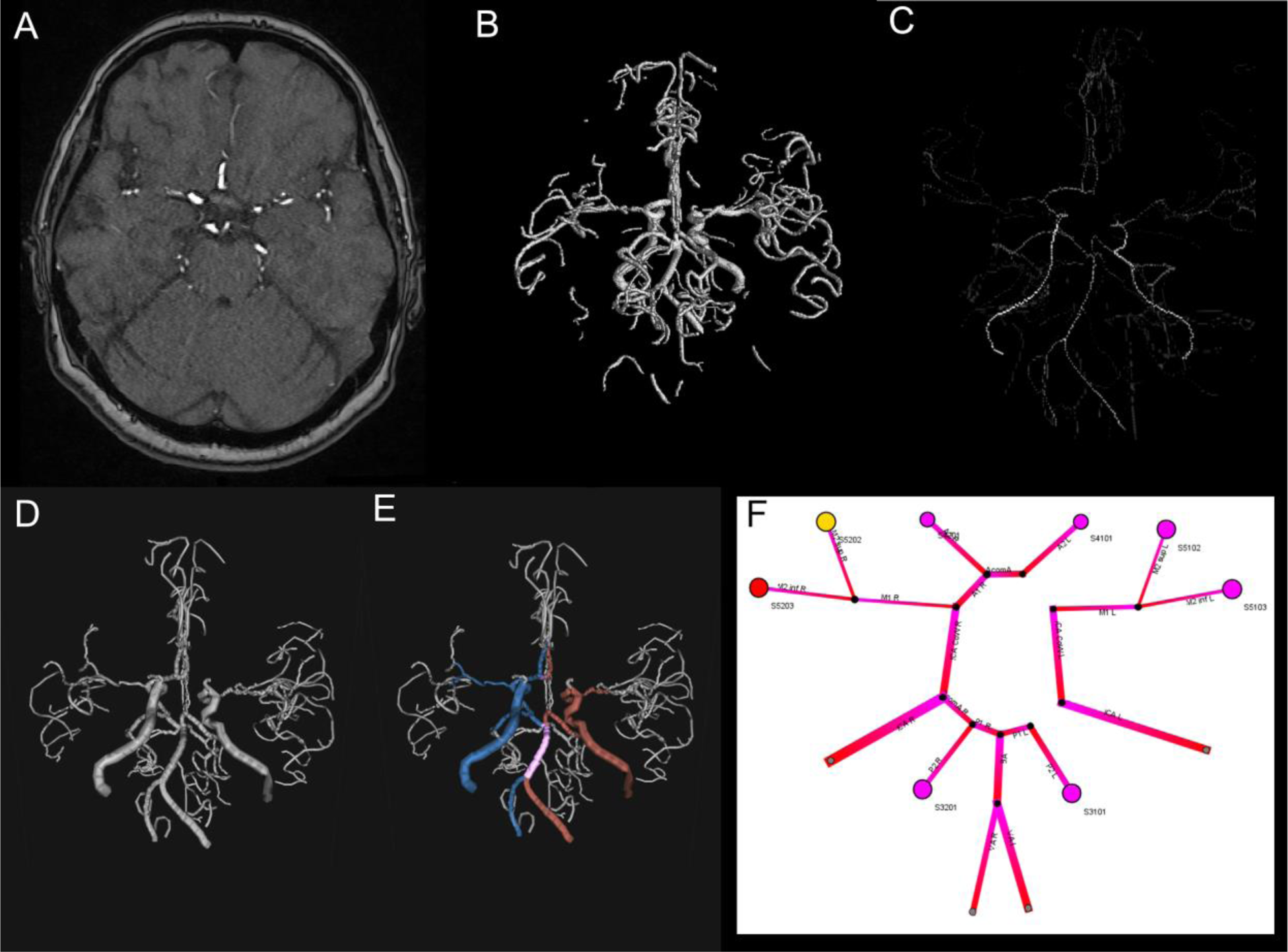
The input is a binary vessel (B) mask segmented from a structural TOF-image(A). This image is then skeletonized (C). This skeleton can be imported in our annotation tool, where the vessel tree is displayed in a 3D fashion and annotation is performed (D and E; red=left, blue=right, purple=median). The annotated vasculature can then be transferred to the simulation tool (F).

**Figure 8:**
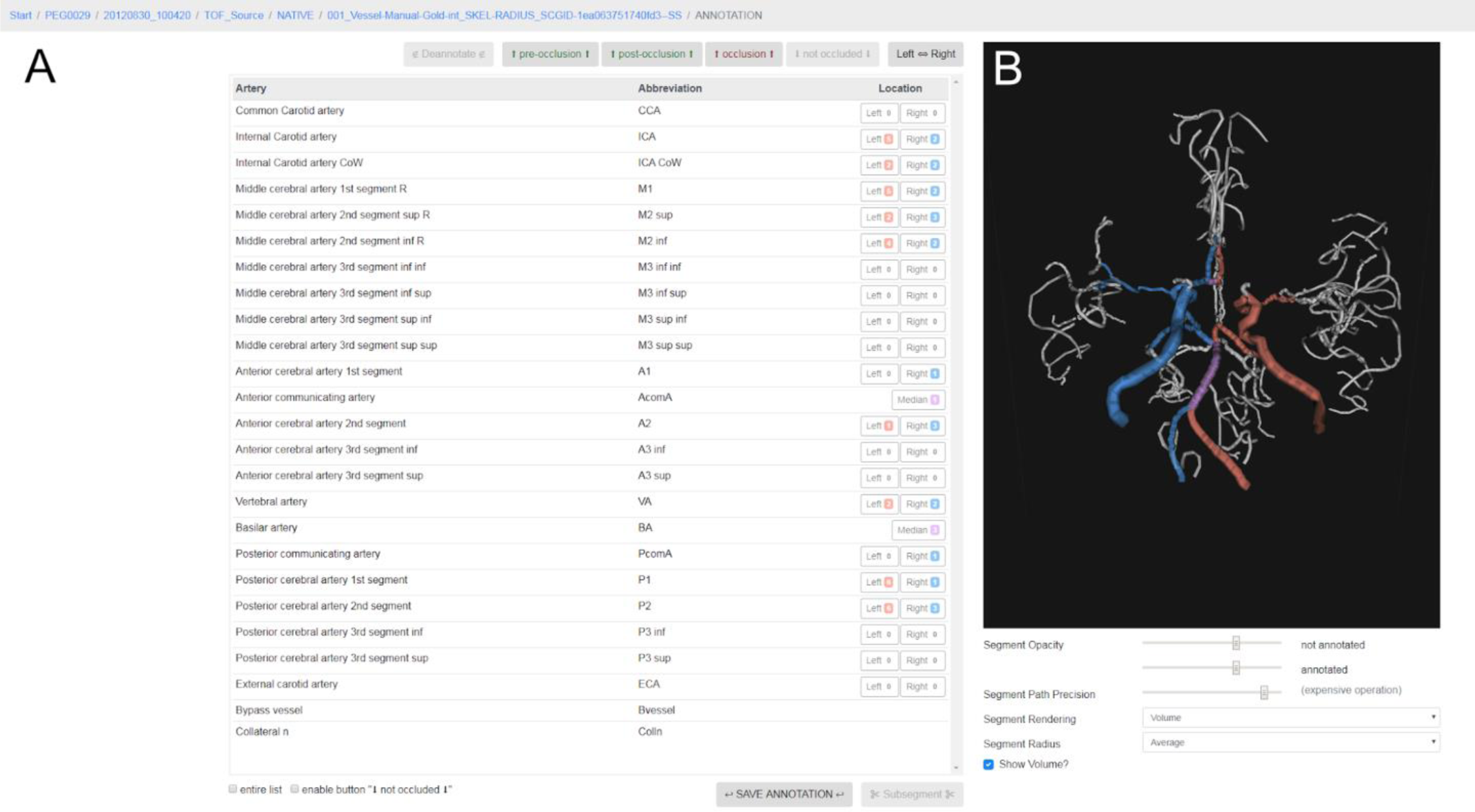
The annotation GUI. It consists of two main components. The segment annotation area, where the vessel segments can be chosen (A) and the 3D view (B), where the imported vessel tree can be manipulated and the vessels can be chosen.

The annotation tool is made up of two main components. The segment annotation area, where the vessel segments can be chosen (Figure 8A) and the 3D view (Figure 8B), where the imported vessel tree can be manipulated.

We implemented 22 segments of brain supplying arteries, namely 3 for the carotid artery (Common, internal and CoW segment), 7 for the MCA (M1, M2 superior, M2 inferior, M3 superior superior, M3 superior inferior, M3 inferior superior, M3 inferior inferior), 5 for the ACA (A1, A2, A3 inferior and A3 superior), 4 for the PCA (P1, P2 and P3 inferior and P3 superior), as well as the Basilar artery, the vertebral artery and the anterior and posterior communicating arteries. It is also possible to add bypass vessels or collateral vessels manually.

Next to segment also the following additional labels can be chosen: “pre-occlusion”, “post-occlusion”, and “occlusion” (the last in case the nature of the occlusion cannot be determined with certainty). To record occlusion next to the type of vessel is important for the following simulation step, as subsequently the simulation will ignore segments with this flag. A video footage of the annotation process was uploaded to zenodo(Frey et al. 2019).

The simulation itself consists of a graphical user interface, that is divided functionally in the toolbar (Figure 9A), the simulation area (Figure 9B), the pressure selection area(9C) and the view selection area (Figure 9D). In the toolbar, simulations can be loaded as well as the type of resistance calculation (here, the default is the resistance calculation presented in the methods section). In C, the pressure boundary conditions can be chosen, for one the blood pressure which determines the driving pressure of the whole system. On the other hand, also the intracranial pressure, which is kept constant for our use case at physiological parameters, but could be increased to simulate conditions with increased intracranial pressure, e.g. in hemorrhagic stroke. In the view selector area, in D, the normal view and the pressure view can be chosen (please see examples in figure 11). Finally, in the simulation area, the individualized simulation of the vasculature can be inspected. Via right click it is possible to derive more information about an edge or a node (Figure 10A and B, respectively). Supply areas are color coded according to the calculated pressure (all values in mmHg: >70 pink, <70 yellow, <60 orange, <50 red). Below 50 mmHg we consider an area being vulnerable to ischemia.

**Figure 9:**
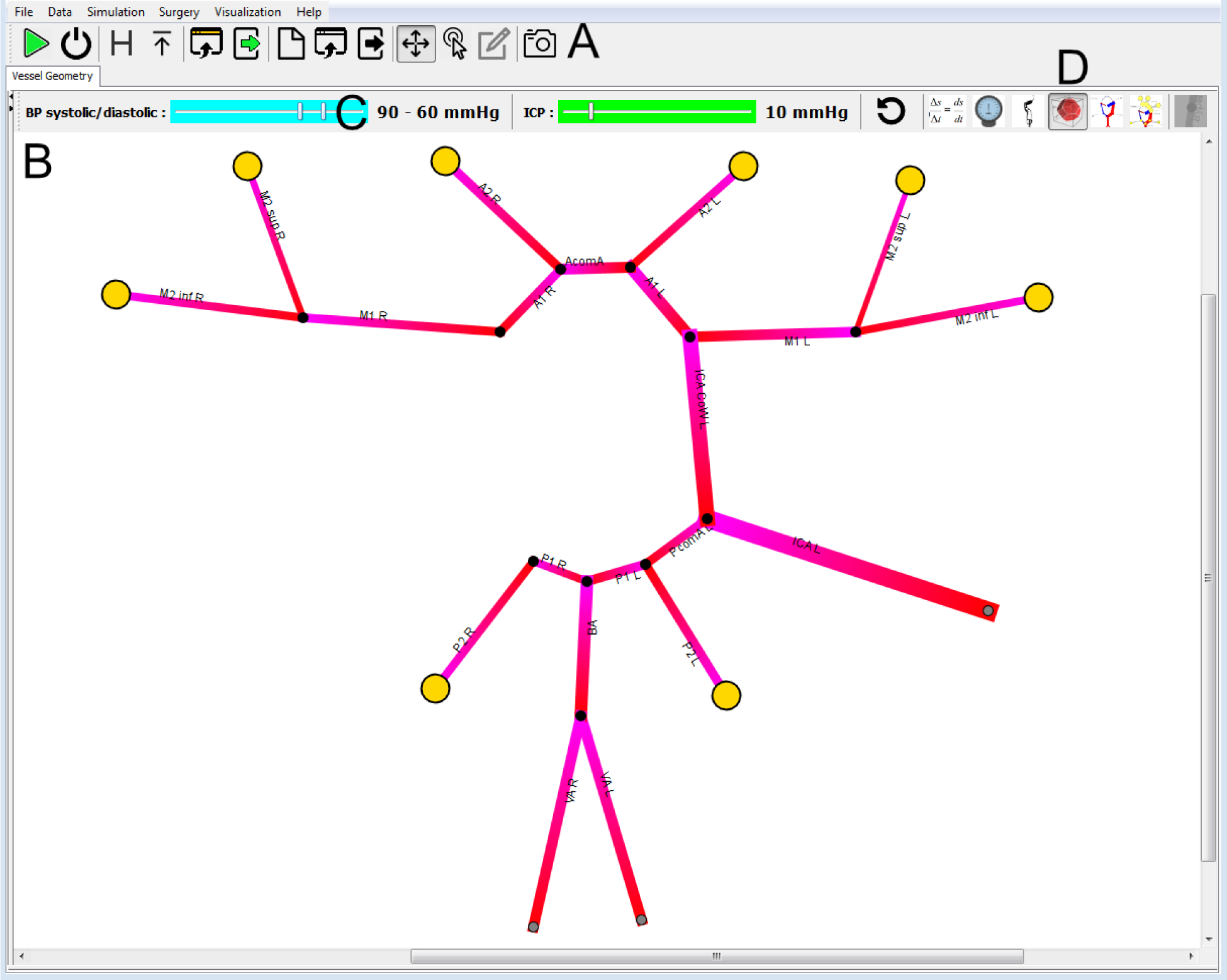
The simulation GUI. It is divided into the toolbar (A), the simulation area (B), the pressure selection area(C) and the view selection area (D).

**Figure 10:**
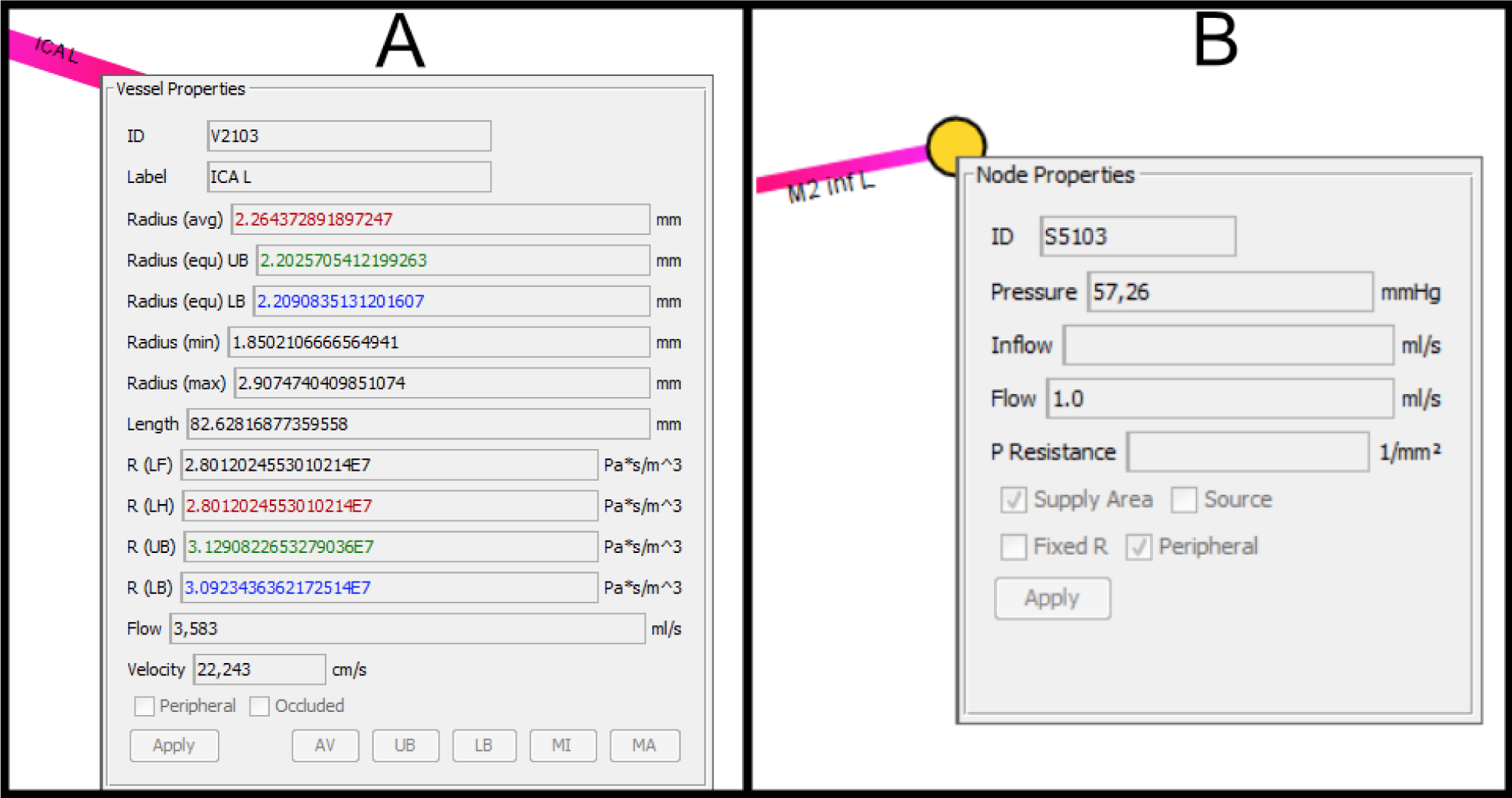
It is possible to inspect additional information for each edge (A) - in this case a left ICA - and each supply area (B).

**Figure 11:**
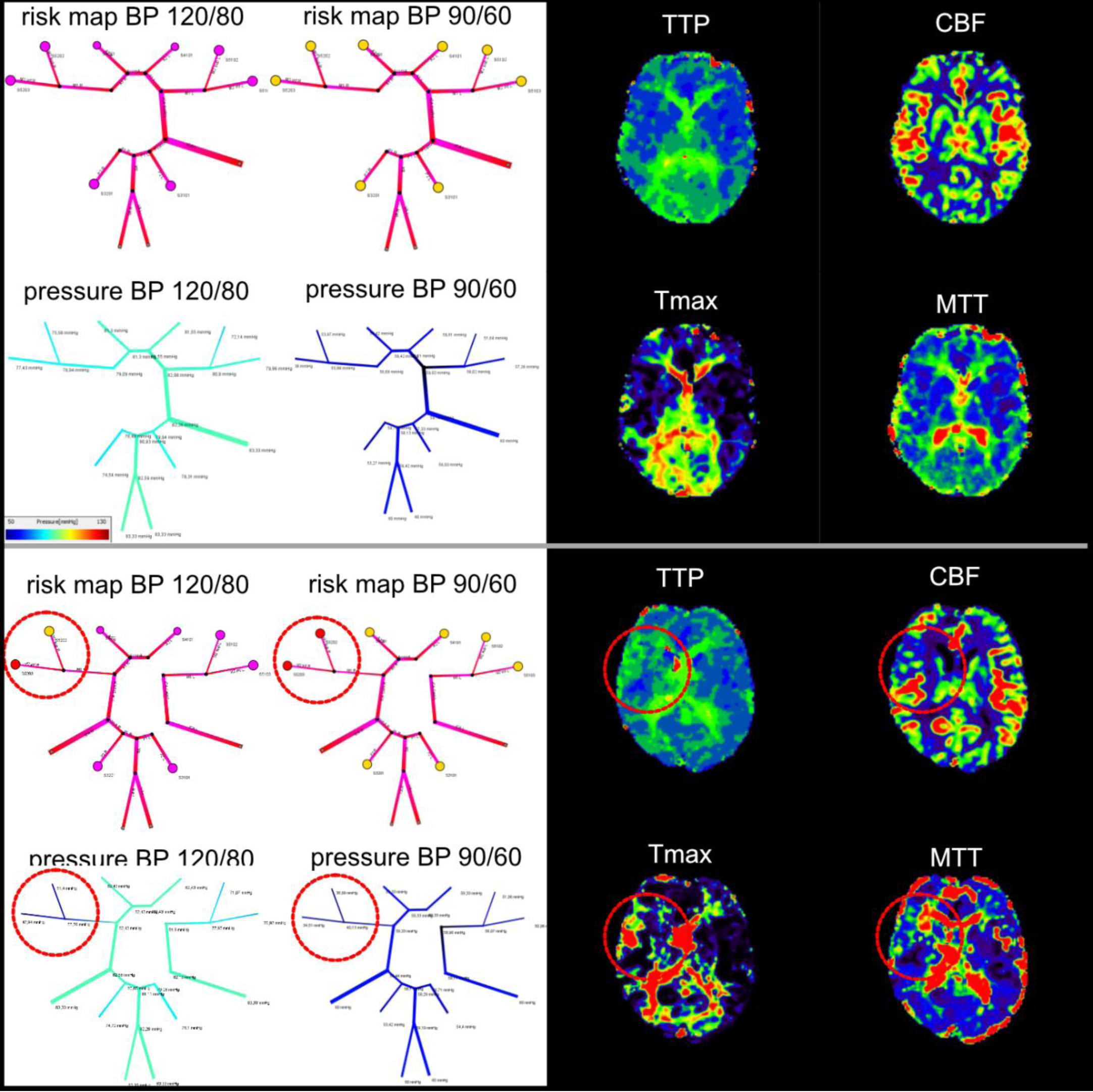
Comparison of simulation results (left) and DSC-imaging (right). In the patient with a proximal common carotid occlusion on the right (upper box), no hemodynamic changes are visible in the DSC perfusion imaging maps indicative of a sufficient CoW to provide all brain areas with sufficient perfusion. The simulation shows correspondingly no areas reaching critical limits, at both normal and lowered blood pressure levels. In the patient with the right MCA stenosis (lower box), the DSC perfusion maps show delay in TPP, MTT and Tmax maps as well as a lower CBF in the right MCA area. This is in line with our simulation which shows critical pressure levels in the M2 inferior node and lowered pressure already at a normal blood pressure of 120/80. When blood pressure is lowered to 90/60, both M2 supply areas become critical in contrast to the other branches. (Supply areas are color coded according to the calculated pressure (all values in mmHg: >70 pink, <70 yellow, <60 orange, <50 red). Below 50 mmHg we consider an area being vulnerable to ischemia.)

### Patient simulations

In the 2 randomly selected patients, we found a high informational overlap between the simulation results and DSC perfusion imaging. In the patient with a proximal common carotid occlusion on the right (figure 11, upper box), no hemodynamic changes are visible in the DSC perfusion imaging maps indicative of a sufficient CoW to provide all brain areas with sufficient perfusion. And indeed, our simulation captures this condition, showing optimal perfusion conditions at normal blood pressure conditions of 120/80 mmHg. Even when lowering the blood pressure to 90/60 mmHg, the pressure levels do not reach critical limits.

This is in contrast to the patient with the right MCA stenosis (figure 11, lower box). The DSC perfusion maps show delay in TPP, MTT and Tmax maps as well as a lower CBF in the right MCA area. This is in line with our simulation which shows critical pressure levels in the M2 inferior node and lowered pressure already at a normal blood pressure of 120/80. When blood pressure is lowered to 90/60, both M2 supply areas become critical in contrast to the other branches. Taken together, both DSC imaging as well as our simulation indicate that patient 2 has an area highly vulnerable to future stroke in the right MCA territory.

### Sensitivity Analysis

An example of the sensitivity analysis is shown in Figure 12, where we give an example how the still normal hemodynamics in a patient with a missing ICA would be affected by additional vessels stenosis. The sensitivity analysis successfully allows individualized simulations of potential disease courses.

**Figure 12:**
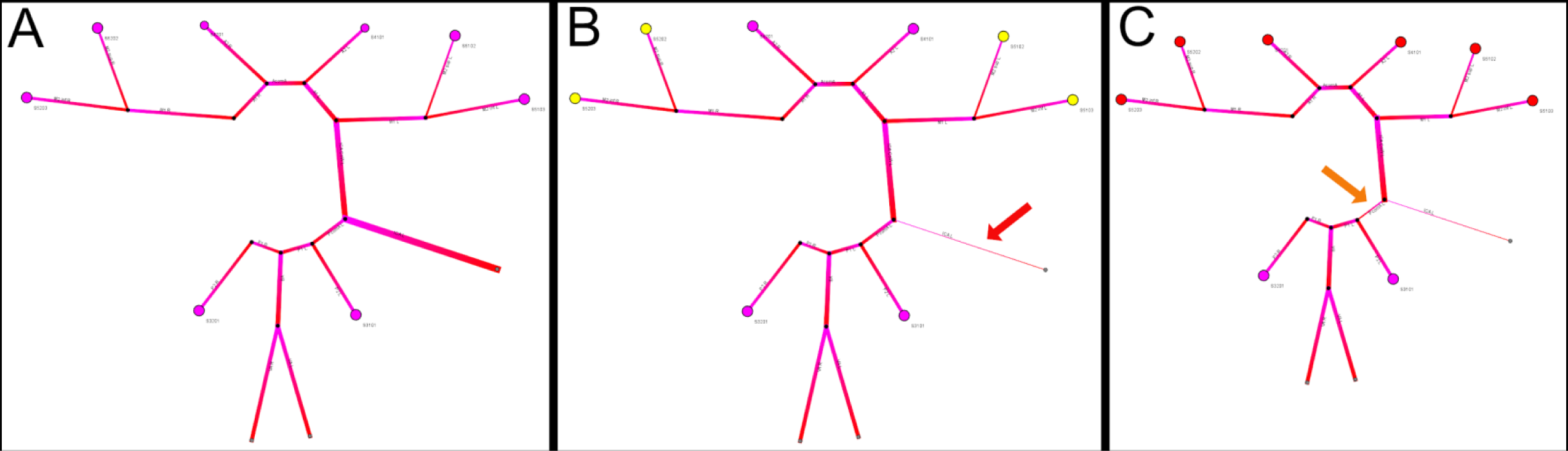
Illustration of the sensitivity analysis. The patient has a right-sided ICA occlusion. All simulation results are calculated under normotension (120/80 mmHg). In the current state (A), the patient has no changes in hemodynamics. Adding a 90% stenosis of the ICA on the other side (B, red arrow) leads to only a little drop in perfusion pressure in both MCA supply areas (yellow supply areas). This can be attributed to a very marked posterior communicating artery on the left side. Adding a 50% stenosis of the left Pcom leads to high vulnerability of the anterior circulation system (red supply areas) of both sides (C, orange arrow).

## Discussion

We present the first precision medicine pipeline for cerebrovascular disease which allows annotation of the arterial vasculature derived from neuroimaging followed by zero-dimensional individualized simulation of brain hemodynamics. Implementation was performed by decreasing the computational burden by a modified MNA, the development of an easy-to-use web user interface-frontend for annotation and a java-based cross-platform simulation tool. Exemplary comparison of our simulation results with DSC-perfusion in two patients with steno-occlusive disease revealed a promising overlap of the provided information. Our results suggest that mechanistic simulation of blood flow derived from routine structural imaging can serve as an individual biomarker for patients with cerebrovascular disease and might be an alternative to complex and potentially harmful perfusion techniques in certain cases.

Stroke is a complex disease with a dynamic progression. The initial infarct area - characterized by rapid neuronal loss- is called the core, which is surrounded by tissue that is slowly surrendering to ischemia, but is still salvageable. The latter area is coined the penumbra and defines the therapeutic target in acute stroke management(Sobesky 2012). For the understanding of stroke and its implications on treatment strategy it is essential that the speed of the penumbra-to-core transformation varies greatly and is highly individual. In particular, for weighing benefit and risk for stroke treatment the high interindividual variance of brain cell death is crucial: Some patients do not have salvageable brain tissue already a few hours after stroke, whereas in others penumbral tissue was found up to 17 hours after stroke (Marchal et al. 1996). This highly individual and variant stroke progression stands in stark contrast to the current “one-size-fits-all” treatment approach in stroke where patients receive treatment based on guidelines usually only within a predefined time window of up to 4.5 hours after stroke(Sobesky 2012; Powers William J. et al. 2018). These time windows were were established by a statistical benefit to risk calculation after lumping together all stroke patients with assumed common pathophysiology. While it is true that there is a net profit for patients when treatment is applied within this time window, it is also clear that many patients do not receive treatment who would benefit from it and at the same time patients receive treatment subjecting them to risk of intervention such as bleeding without its benefit. This is due to the above-mentioned fact that, in reality, several stroke subpopulations exist. Here, precision medicine accounts for the individual features and will improve outcome by personalizing treatment (Livne 2019). Precision medicine utilizes mathematical techniques and available digital data to provide individualized predictions for patients(Hinman et al. 2017; Rostanski and Marshall 2016). It relies on the presence of informative data allowing the differentiation of pathology patterns(Hinman et al. 2017). In stroke, it has been shown that measuring the penumbra through perfusion as a surrogate is one of the most promising approaches(Sobesky 2012; Campbell et al. 2019). And indeed, perfusion imaging-based selection for treatment beyond the established time windows is an evidenced precision medicine approach in stroke(Campbell et al. 2019). For the selection process, predictive modelling might be also applicable, see e.g. Livne et al(Livne et al. 2018). A drawback of this approach, however, is the application of perfusion measurement techniques which are potentially harmful through contrast agents, inevitably prolong the imaging time and are problematic to standardize across centers(Sobesky 2012; Calamante 2013; Zaharchuk 2011). Similar considerations apply to chronic steno-occlusive disease. These are patients with continuously worsening symptoms of atherosclerosis which have a high likelihood for a future stroke event. In these patients, potentially harmful perfusion imaging techniques should not be used a priori. Contrast-agent free perfusion imaging methods can be used, but are - as mentioned above - hard to standardize. Thus, alternatives are warranted and mechanistic simulations are promising methods. Here, the relevant (patho)physiological biomarker is not directly measured, but mathematically inferred from conditions recorded through other measurements.

As suggested by our work, in the case of cerebrovascular disease, we can infer information about hemodynamics from the individual vasculature of a given patient. We successfully built a pipeline that can extract the vessel information, allows annotation of the vessels and simulates hemodynamic information which we were able to relate to clinical DSC-perfusion imaging. While these results are - naturally - only preliminary and need validation, they pave the way for further development of techniques that might make the need for perfusion imaging in cerebrovascular disease obsolete for some patients while still providing the necessary information for precision medicine selection of patients for prevention and treatment.

Given that mechanistic simulations work on a priori assumptions about the biological system and perfusion measurements actually record dynamic information, it is unlikely that the information provided by both systems will always be a complete match. Our results suggest, however, that the information might be intersecting enough to allow treatment relevant predictions on a patient level, and consequently to avoid harmful imaging procedures. Thus, when the question is for example about a general status, e.g. “is there a general vulnerability in the right MCA area for ischemia”, mechanistic simulations might be able to provide this information instead of direct perfusion measurements. Also, since many variants of the CoW exist, the simulation could allow the identification of patients with high-risk for stroke owing to their individual CoW configuration(Pascalau et al. 2018).

Another potentially big advantage of mechanistic simulations is the possibility to simulate interventions. In chronic steno-occlusive disease, like carotid stenosis or Moya-Moya disease, potential lumen reopening interventions or EC-IC bypass surgery can be performed. With our solution as presented in this work, it would be feasible to simulate the reopening of a vessel and thus simulate the post-intervention status.

With our framework, it is possible to simulate the response of the vasculature to changes in blood pressure. This can potentially be highly important not only for the determination of areas-at-risk for ischemia but also to predict the response to interventions and surgery, e.g. blood pressure drops during surgery. This is not possible with perfusion measurements, which can only provide a snap-shot of the status-quo. Approximations can be done with acetazolamide challenge measurement(Yen et al. 2002; Ma et al. 2007), but this requires repeated measurements, the application of a drug, and can only be performed within the physiologically tolerable range. A clear advantage of direct dynamic perfusion measurements, on the other hand, is with high likelihood still the recording of subtle changes and small lesions. Importantly, we thus do not claim that mechanistic modeling might be able to make all perfusion measurements obsolete.

As a major limitation of our work, it is clear that our results are exploratory and hypothesis-generating(Kimmelman, Mogil, and Dirnagl 2014). While the exemplary assessment of two randomly selected patients with cerebrovascular disease showed promising and encouraging results, a systematic validation was beyond the scope of the current work, but will be performed in the future. We believe, however, that our results are motivating to boost the translation of the work done in the past on the development of mechanistic modeling of hemodynamics into the clinical setting. We implemented a 0-dimensional (D) model of hemodynamics which exploits the similarities of such a network to an electric circuit. Next to these 1D and 3D models exist, for an overview of existing methods see Perera et al and Leguy et al(Perera 2019; Leguy 2019). While 1D and 3D models are more suited to model local changes, 0D models are more suited to model the general vasculature, but they can be combined to provide complementary information(Chau and Ho 2020). Here, our framework builds on existing work, but adds A) the calculation of the resistance over segments with variable diameters. B) an easy to code adjusted implementation of the modified nodal analysis which reduces computational demands and C) a graphical user interface tailored for inspection and manipulation of the simulation. This facilitates the application of such a framework, which is promising as there is much promise in mechanistic modeling of blood flow and perfusion for clinical applications in cerebrovascular disease. There is, however, a need to personalize these approaches. Here, we present the first precision medicine pipeline that allows annotation and then individualized simulation based on structural vessel imaging. This work paves the way for future personalized applications. Another major consideration is collateral flow which is defined as functional and anatomical enlargement of smaller vessels supplying the same area when a major vessel pathway is blocked. On one hand, a limitation of the current version of the framework is the lack of collateral flow beyond CoW collaterals. And indeed, for certain use cases this might be crucial, for example for bypass surgery. On the other hand, however, especially for the use case at hand - identification of areas at risk in cerebrovascular disease - this might not be necessary. Let us assume that a hypothetical MCA area downstream to an MCA occlusion is flagged by the simulation framework as being at risk. In reality, the cerebral blood flow of that area might be unchanged owing to leptomeningeal collaterals. However, maps indicating delayed arrival like TTP and Tmax would still flag that area also in DSC imaging as vulnerable to an ischemic event showing hyperintense areas indicative of delay. This means that while the calculated pressure threshold for that area would probably be too low in our simulation, they would still indicate a vulnerable area correctly. This situation would be highly similar to the transit delay artifact in arterial spin labeling (ASL). Here, the delayed flow leads to an accumulation of labeled spins within large arteries leading to hyperintense perfusion values where normal or lower blood flow was expected(Mutke et al. 2014). However, it was shown that this hyperintense artifact corresponds to TTP and Tmax delays and thus might be used to identify vulnerable areas(Mutke et al. 2014; Amukotuwa, Yu, and Zaharchuk 2016). Importantly, however, these considerations are subject to thorough clinical validation which we will perform in the future.

Our work has other limitations. First, the simulation values were not compared to a gold-standard. Importantly, however, it is very difficult to derive *individualized* gold-standard values for arterial flow. Other noninvasive methods are either non-goldstandards themselves, like MRI flow measurement methods, and/or cannot access the complete vasculature, like Doppler- sonography. Intra-operative direct vessel flow measurements recorded during EC-IC bypass surgery might be an option for gold-standard measurements - which we will explore in the future -, but were not available for the current study. Second, we would like to point out that all relevant pre-processing steps in the pipeline - segmentation, skeletonization, and annotation - are done currently manually in the pipeline. With the advent of powerful machine learning segmentation methods in recent years, it is very likely that these steps can be automated with sufficient performance. Our group has just recently presented a deep learning method to segment the vasculature from structural scans with very high accuracy(Livne et al. 2019). The application of deep learning for skeletonization and also automated annotation is a current focus of our group. We are thus confident that for future potential applications in the clinical setting simulation results will be obtainable in real-time, at the scanner console, in a few years.

## Conclusion

We present the first precision medicine pipeline for cerebrovascular disease which allows annotation of the arterial vasculature derived from structural vessel imaging followed by personalized simulation of brain hemodynamics. This paves the way for further development of precision medicine in stroke using novel biomarkers and might make the application of harmful and complex perfusion methods obsolete for certain use cases.

## Data Availability

Due to data privacy laws patient data cannot be made accessible.

https://doi.org/10.5281/zenodo.3576353.

## Appendix 1

**Derivation of extrapolated resistance**

The following details the derivation of *R*_*ext*_ as presented in formula 4.

As a first intuitive replacement, the radius *r*(*l*) should be replaced by a function that linearly connects the start-voxel *i* with the end-voxel *i* + 1. This allows to yield the function in the real number space ℕ such that the integral in the resistance equation (equation 3) can be calculated. The function of the radius *r* over a segment *l* can be written as:

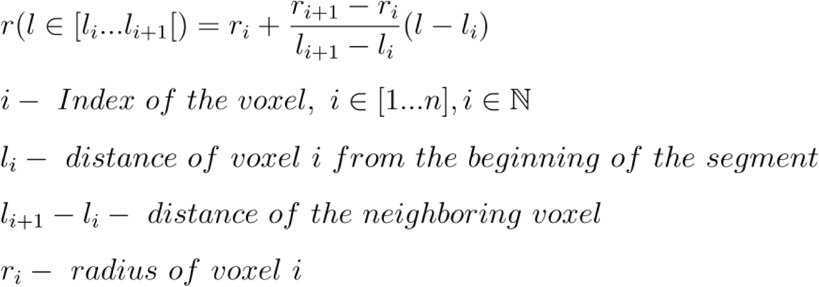

An integral can be formed for all functions in the real number range. However, a primitive function can only be determined for continuously differentiable sections. Therefore it is necessary to solve the problem analytically. The positions where the function can not be differentiated are thus handled as follows: The sum rule makes it possible to divide the total integral into integrals over the subsegments whose integral boundaries lie exactly in the points of discontinuity. The sum of the integrals over the subsegments then provides the total integral:

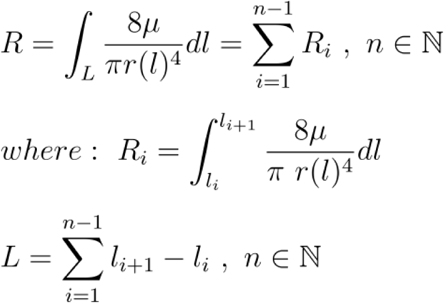

Here the distance between the voxels are considered as subsegments of the overall segment. A full vessel therefore consists of *n* voxels and thus of *n* − 1 subsegments. The integral over the subsegments is defined as:

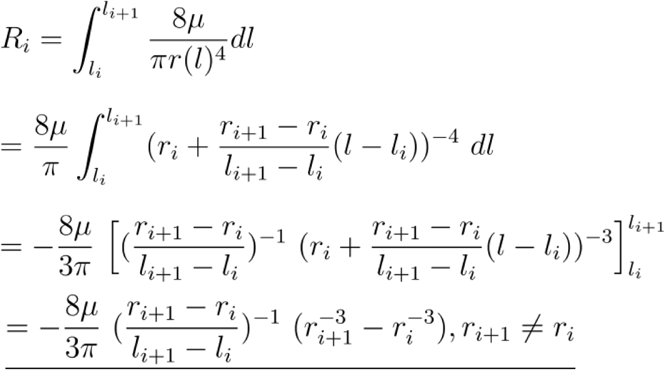

For the case of *r*_i+1_ = *r*_i_ the integral on the subsegment is simplified to:

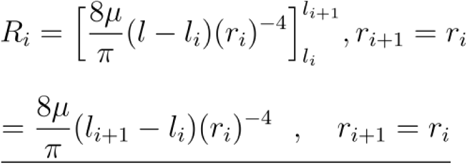

This finally results in the total integral as depicted by equation 4:

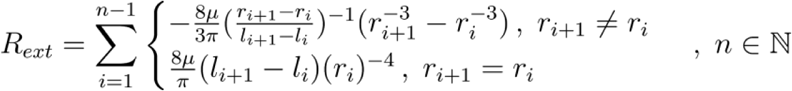

## Appendix 2: Physiological data

The information in table 2 below was used for the generation of a normal vessel tree in the absence of individual patient data. The data is taken from the publication of Alastruey et al.(Alastruey et al. 2007) who aggregated physiological data collected by Stergiopulos et al.(Stergiopulos, Young, and Rogge 1992), Fahrig et al.(Fahrig et al. 1999) and Moore et al.(Moore et al. 2006). In the table, we have listed the segments just once under the presumption of symmetry between the left and right hemispheres of the brain.

**Table 2:** Physiological data used in the model[5].

| Arterial segment | Length[mm] | Radius[mm] |
| --- | --- | --- |
| In. Carotid I (L/R) | 177 | 2.00 |
| Vertebral (L/R) | 148 | 1.36 |
| PcomA (L/R) | 15 | 0.73 |
| M1 (L/R) | 119 | 1.43 |
| A1 (L/R) | 12 | 1.17 |
| A2 (L/R) | 103 | 1.20 |
| P1 (L/R) | 5 | 1.07 |
| P2 (L/R) | 86 | 1.05 |
| AcomA | 3 | 0.74 |
| Basilar | 29 | 1.62 |

This data together with the planar graph depicted by figure 1, was used to build our model of the circle of Willis. When patient data is imported these values are replaced by the individual patient data.

## Acknowledgments

We acknowledge Kim Mouridsen and Mikkel Bo Hansen from the Centre for Functionally Integrative Neuroscience(CFIN) from Aarhus University, Denmark, for providing the pgui perfusion software (V1.0) for research purposes.

